# Novel markers of cognitive impairment and resilience using digital and plasma biomarkers across different neurodegenerative diseases

**DOI:** 10.1101/2025.11.13.25340143

**Authors:** S Toniolo, S Zhao, M Broulidakis, C Gendarini, D Sinha, N Tahira, A Scholcz, B Amein, A Fower, C van Duijn, S Thompson, S Manohar, I Koychev, M Husain

## Abstract

Blood-based biomarkers enable early diagnosis of neurodegenerative diseases in a cost-effective and non-invasive way. Digital cognitive measures can capture early signs of cognitive impairment remotely and at scale. Their combination offers important opportunities for effective screening. We evaluated the performance of a new proteomic assay and a fully remote digital platform in a large memory clinic population (n = 391). Plasma biomarkers were measured using the NUcleic acid Linked Immuno-Sandwich Assay (NULISA) central nervous system panel. OCTAL (Oxford Cognitive Testing Portal) digital cognitive testing platform was used to measure cognition. Distinct proteomic patterns could be identified across different neurodegenerative diseases and associate with specific cognitive functions. Alzheimer’s Disease (e.g., pTau217), and novel proteomics biomarkers (e.g., ACHE and IL6R) were highly correlated with worse cognition. A cluster of biomarkers, including PRDX6, was overexpressed in healthy controls and associated with better cognition, indexing cognitive resilience.

## Introduction

Since the introduction of the ATN criteria in 2018^1^, the definition of Alzheimer’s disease (AD) has shifted from a clinical syndrome to a biological construct, whereby the presence of a multidomain cognitive impairment is neither necessary nor sufficient to define the disease.

Plasma biomarkers have emerged as sensitive and scalable tools to measure disease-specific biological pathways, with the advantage of being more cost-effective and less invasive than gold-standard measures such as cerebrospinal fluid analysis (CSF) and positron emission tomography (PET)^2^. Currently, phosphorylated tau species, particularly phosphorylated tau (pTau)217, are arguably the best plasma biomarkers available to detect AD pathology in vivo ^3,4^. Whilst the field has been fuelled primarily by refining methods to detect AD pathology, understanding the contribution of non-AD pathology is essential, since co-pathology, especially at older age, is the norm rather than the exception and has been shown to have an additive detrimental effect on cognition^5^. The new AD criteria of 2023 have acknowledged such complexity, with the introduction of a 2D system, where cognitive impairment and biomarker abnormalities constitute two axes of a variation ^6^. This marks an important step forward. However, whilst factors such as alpha-synuclein^7^, TDP-43 pathology^8^, and vascular burden^9^ are known to exacerbate cognitive impairment, biological markers related to cognitive resilience have, to date, remained elusive.

Proteomics is becoming more widely adopted to explore such complexities, and has been given additional momentum by large scale studies such as the Global Neurodegeneration Proteomics Consortium (GNPC)^10^, using SOMAmer technology. However, it lacks clinically useful markers in AD such as pTau217 and is expensive to run. Nucleic acid-Linked Immuno-Sandwich Assay (NULISA), is a novel proteomic platform which enables measurement of many biomarkers in plasma and serum with attomolar sensitivity^11^. The NULISAseq central nervous system (CNS) disease panel analyses over 120 proteins linked to different neurodegenerative diseases, including but not limited to AD biomarkers. This open the door to both biological characterization of clinical phenotypes and exploration of novel biomarkers within a single analysis. Recent data supports its use in AD^12,13,14^ as well as across different neurodegenerative diseases, including Lewy body disease (DLB) and Frontotemporal dementia (FTD)^13,15^. However, most studies are limited to comparisons between two diagnostic groups, with limited transdiagnostic generalizability, or do not explore the impact on cognition.

Biomarker testing in clinical practice is currently recommended only in patients with cognitive impairment ^6,16,17,18^. This is because pretest probabilities vary by cognitive stage, with patients with subjective cognitive decline (SCD) having higher false positive rates compared to patients with objective cognitive impairment^19^. Moreover, only patients with objective cognitive impairment are eligible for Aβ-lowering immunotherapies at present ^20,21^. Therefore, identifying, at scale, people with objective cognitive impairment remains essential and complementary to studying their biological risk of carrying AD pathology.

In parallel with the plasma biomarkers revolution, the field of cognitive testing has also undergone a radical transformation. Digital platforms are becoming more widely adopted given the lower administration times, hence lower burden on the healthcare system, and wider scalability for the purpose of screening, diagnosis and follow-up^22,23^. OCTAL (Oxford Cognitive Testing Portal) is a fully remote platform, which can discriminate effectively between healthy controls and patients with AD dementia^24,25^. It shows high test-retest reliability and cross-cultural applicability^24^. Performance on several of its digital tests correlate closely with core AD biomarkers, particularly pTau species^25^.

From the OCTAL test battery, two tests have been deployed for use in the current study: the Oxford Memory Test (OMT) and a digital version of the Trail Making Test (TMT). OMT is a visual short-term memory task, which has been extensively used across different neurological conditions, including SCD, mild cognitive impairment (MCI) and AD dementia^26,27,28,29,30,31,32,33^. Performance on this test can discriminate effectively between patients with AD dementia and healthy individuals, both in its tablet version and fully remote online version ^24,25,29^. It has been shown to be able to capture early signs of cognitive decline alongside the AD continuum, as in patients with SCD and MCI ^24,25,29^, to track disease progression longitudinally, and to predict hippocampal atrophy as well as standard neuropsychological tests^24,25,29^. The digital version of OMT has not been used by patients with other primary dementias such as DLB, FTD and Corticobasal syndrome (CBS) prior to this study. No studies have explored the relationship of NULISA’s markers and performance on digital cognitive testing.

In this study the relationship between plasma biomarkers and digital testing using OCTAL was investigated through linear partial and canonical correlations in a large memory cohort, encompassing patients with SCD, MCI, AD, DLB, FTD and CBS as well as cognitively healthy controls. A network analysis was performed to understand the latent structure connecting different biomarkers. Models’ performance using digital measures, biological measures and their combination were tested to measure classification accuracy in discriminating between healthy subjects and patients with AD and other dementias. Finally, biomarkers levels were compared across different neurodegenerative diseases.

## Results

### PCA of plasma biomarkers

Given the high dimensionality of the biomarkers and behavioural data, we independently applied a dimensionality reduction technique, e.g., Principal Components Analysis (PCA), on both sets of variables. The PCA of plasma biomarkers yielded 10 components (**Figure 1**, Extended data).

**Figure 1.**
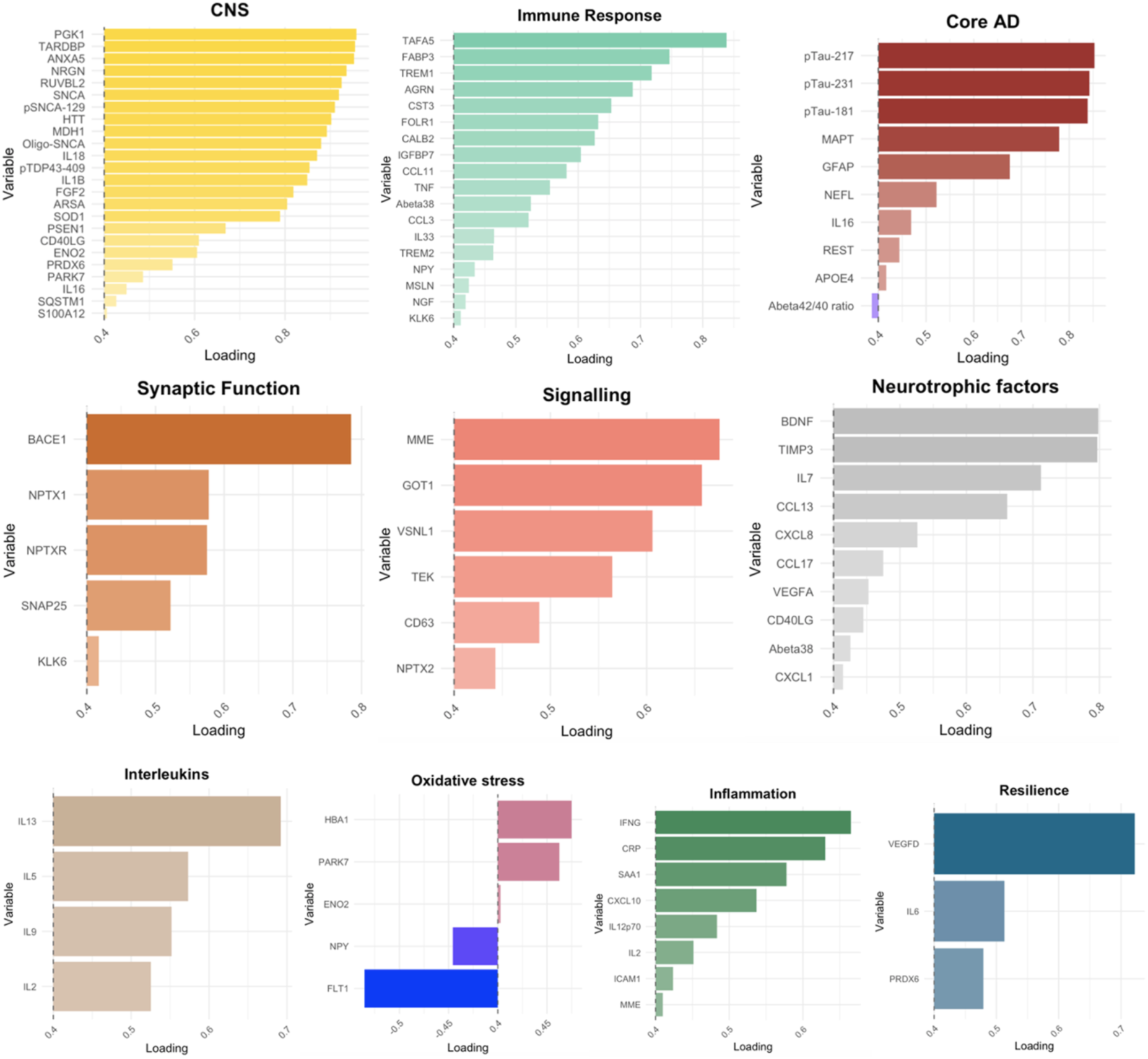
Principal component analysis: Component loadings for biomarkers. On the y-axes, biomarkers included in each PC component. CNS = Central nervous system. On the x-axis, factor loading for each component. Only loadings > 0.4 were retained. Stronger loadings are represented by darker colours and longer bars.

### PCA of digital cognitive assessment

The PCA on the digital cognitive metrics yielded 5 PC components. The first component, ***Spatial precision***, included Absolute Error, Imprecision and Localization Time from the OMT task. Misbinding and Guessing (OMT) constituted the second component, ***Spatial identity***. The third component, ***Executive functions***, was represented by the TMT task. The fourth component, ***Reaction times***, included Identification and Localization Time (OMT), whilst the fifth component, ***Accuracy***, was represented by Identification Accuracy (OMT). Loadings of each component can be found in Extended data.

### Correlation between digital and biomarker PCs

We next investigated the relationship between PCs derived from biomarkers and digital measures. PC3 (*Core AD*), showed the highest positive association with cognitive performance (**Figure 2**), where the higher the biomarkers’ values the worse the cognition, with the exception of the Aβ42/40 ratio which had the opposite direction. This was true across the first four digital PCs, namely *Spatial precision*, *Spatial identity*, *Executive functions*, and *Reaction times.* PC9 (*Inflammation*), was also positively associated with cognitive performance, across *Spatial precision*, *Spatial identity*, *Executive functions*, but not *Reaction times* PCs, and was negatively associated with *Accuracy*, where the higher the inflammatory biomarkers levels, the worse the accuracy at the OMT task. PC5 (*Signalling*) also showed a positive correlation with the *Spatial precision* PC. Conversely, PC1 (*CNS)*, PC4 (*Synaptic function)* and PC10 (*Resilience)* showed negative correlations with cognitive performance, where the higher the values, the better the cognitive performance.

**Figure 2.**
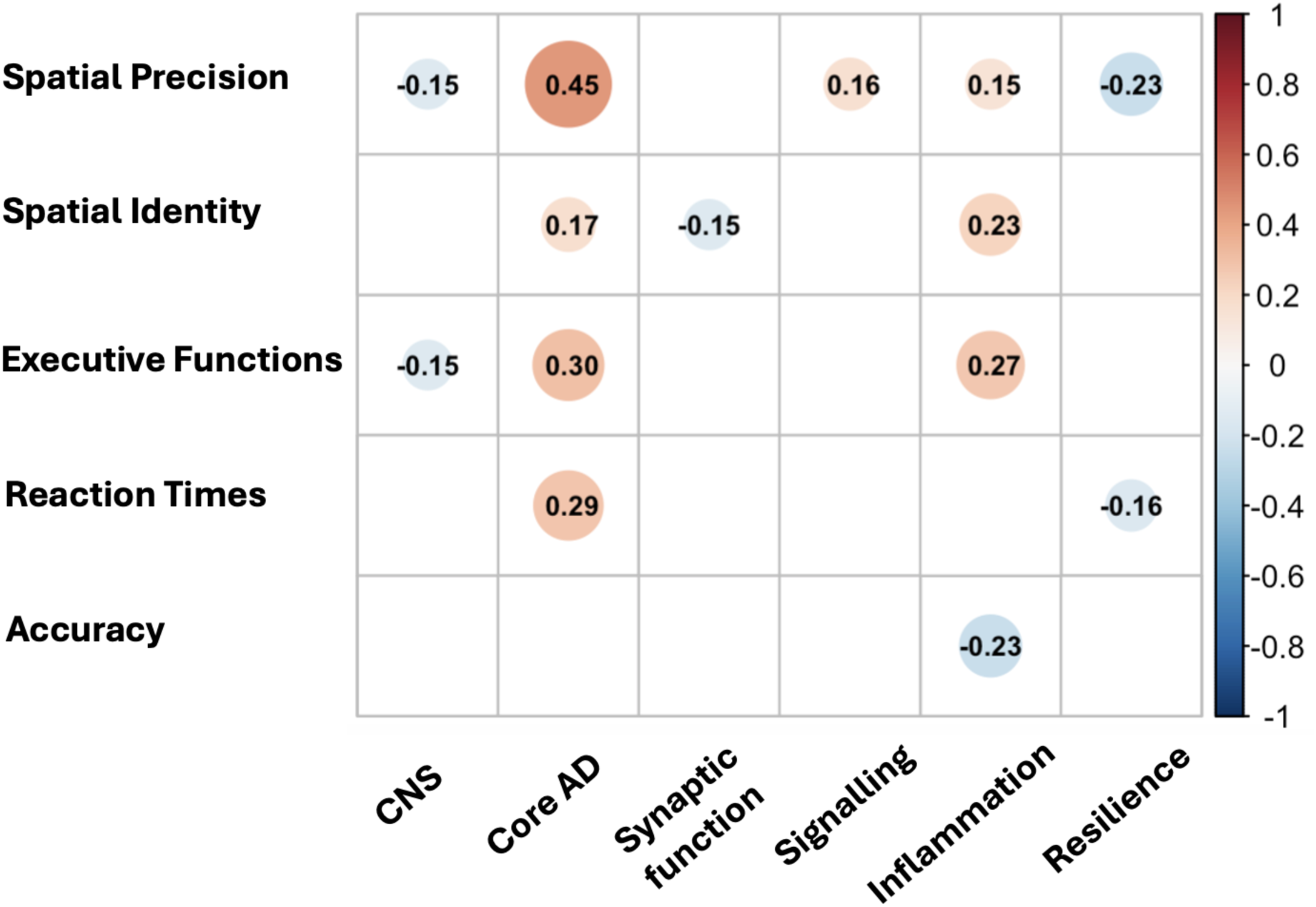
Correlation between digital metrics and plasma biomarkers’ PCs. Spearman rank test, using age, sex, and education as covariates was used to compute correlation between PCs. The Benjamini–Hochberg method, which controls the false discovery rate (FDR), was applied to correct for multiple comparisons. Only statistically significant correlations are shown. Warmer colours represent positive correlations, whilst colder colours represent negative correlations.

### Canonical correlation between digital and plasma biomarkers

Whilst using dimensionality reduction can be useful in clustering variables within a class and reducing multiple comparisons when correlating these to another set of variables, it fails to capture global associations between two different classes. Canonical correlation analysis finds linear combinations of two sets of variables, in this case digital metrics and plasma biomarkers, that have a maximum correlation with each other ^40^. This is achieved without prior intra-class clustering using a PC approach. Instead, it constructs pairs of canonical components, each being a linear combination of the variables within one class, so that the correlation with the other class is maximized. In this study, the canonical correlation analysis yielded 4 components, and only the first one (strongest association) was retained. This revealed a strong association (0.742) between digital measures and a subset of biomarkers, spanning across different previously defined PCs (**Figure 3**). Some of the top plasma biomarkers belonged to PC3 (Core AD), including but not limited to pTau species (pTau217, pTau231, pTau181), as well as glial acidic fibrillary protein (GFAP), neurofilament light chain (NEFL), and microtubule-associated protein tau (MAPT). Other proteins not belonging to the Core AD cluster, such as Acetylcholinesterase (ACHE), Interleukin-6 receptor (IL6R) and several other proteins were also strongly associated with cognition. Notably, Peroxiredoxin-6 (PRDX6) had a strong negative loading, suggesting a protective role on cognition.

**Figure 3.**
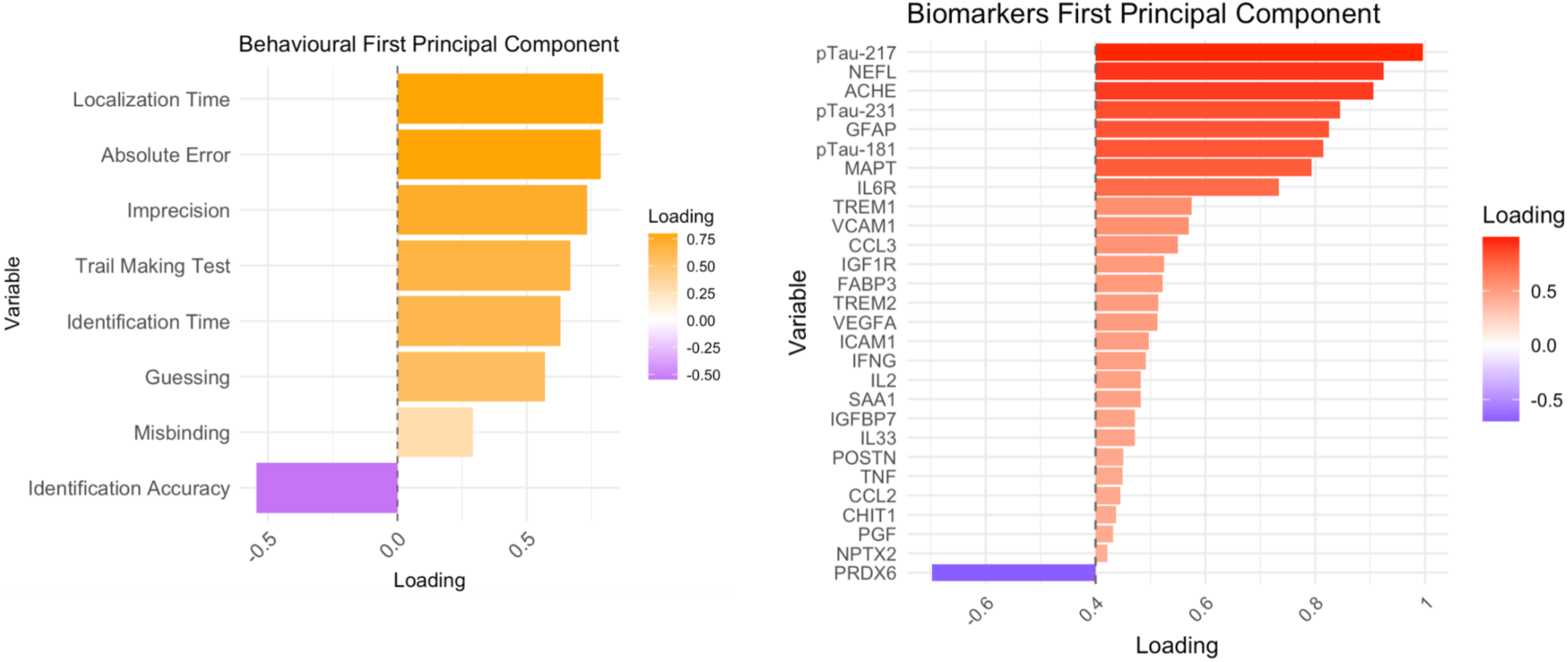
Canonical correlation analysis: factor loadings for behavioural metrics and biomarkers. Warmer colours (orange and red) represent positive correlations, whilst colder colours (violet) represent negative correlations. Stronger loadings are represented by darker colours and longer bars. For biomarkers, only factor loadings > 0.4 were retained.

The top positive metrics for the behavioural component belonged to the *Spatial Precision* behavioural PC. The negative loading within the digital measures represents Identification accuracy, i.e. the lower the accuracy, the higher the biomarkers levels.

### Model comparisons for patient classification

As next step, the accuracy of behavioural measures and plasma biomarkers, alone or in combination, was tested in discriminating between diagnostic groups. These were HC, AD and non-AD dementias (DLB, FTD and CBS).

In the comparison between AD and HC, the best behavioural model (‘Behavioural’), consisted of a combination of TMT (β = 3.91, p < 0.001), Absolute Error (OMT) (β = 0.96, p = 0.022), and Identification Accuracy (OMT) (β =-0.80, p = 0.001), with an AUC of 0.933 (confidence interval (CI) 0.88-0.98), (**Figure 4**, panel a and Extended data). When biomarkers were used as predictors for group discrimination, the winning model (‘Biomarkers’) consisted of a combination of pTau217 (β = 2.82, p < 0.001), ACHE (β = 1.89, p < 0.001), and PRDX6 (β =-0.67, p = 0.006), with an AUC of 0.966 (CI 0.93-1). As a comparison, a model with pTau217 alone (‘pTau217 alone’) had an AUC of 0.940 (CI 0.897-0.983). The combination of the best behavioural metrics and the best biomarkers model (‘Combined’) had an AUC of 0.99 (CI 0.98-1). When comparing models, accuracy of the ‘Behavioural’ model was not statistically different compared to the ‘Biomarkers’ model (χ² = 0.857, p-value = 0.354), or the ‘pTau217 alone’ model (χ² = 1.88, p-value = 0.170 and the ‘Biomarkers’ model was not superior to ‘pTau217 alone’ (χ² = 0.6, p-value = 0.439). However, the ‘Combined’ model was better than all the other models (‘Behavioural’ model (χ² = 9.965, p-value = 0.002), the Biomarkers model (χ² = 9.301, p-value = 0.002), and ‘pTau217 alone’ (χ² test = 4, p-value = 0.045)), (**Figure 4**, panel b and Extended data).

**Figure 4.**
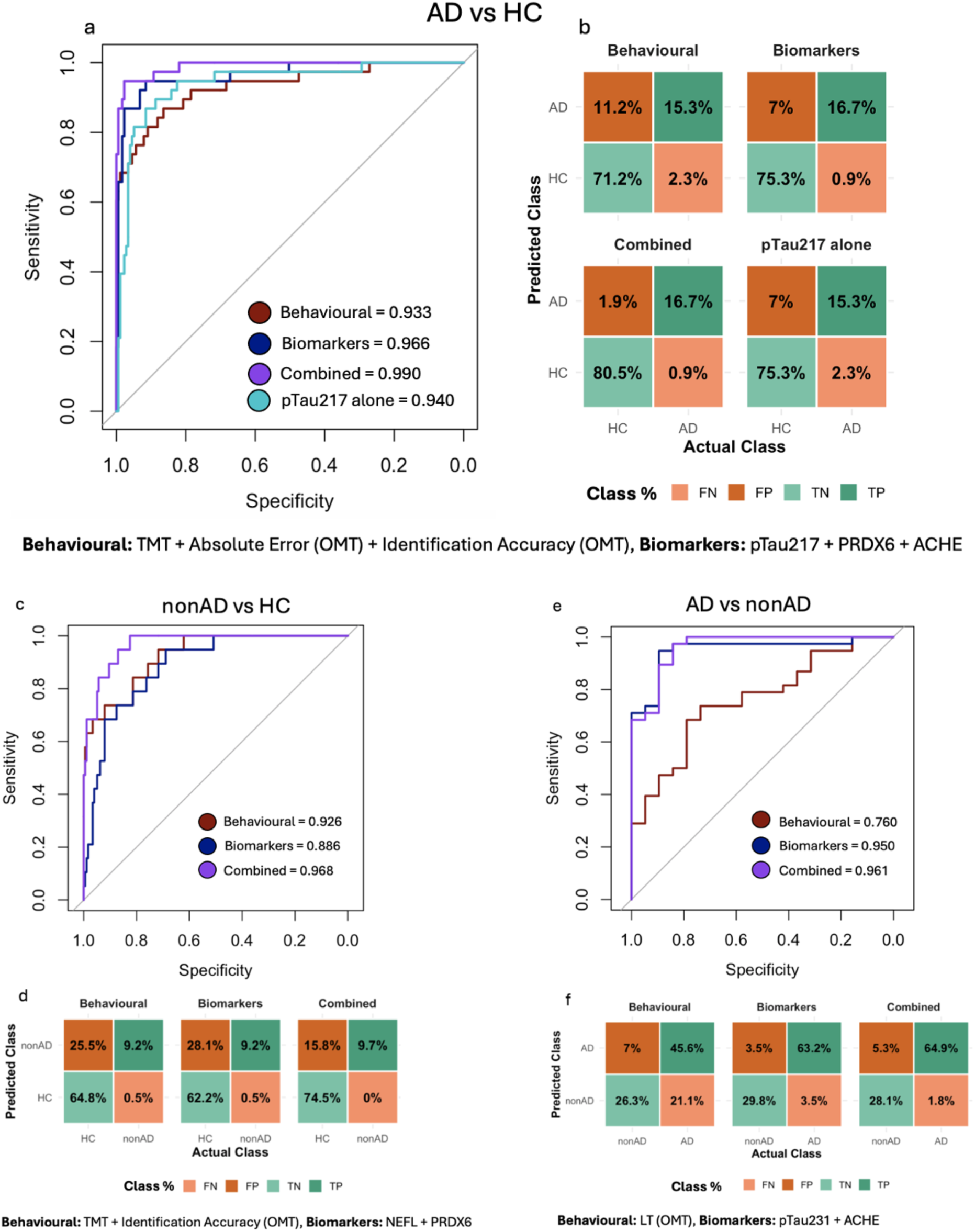
Model comparisons across behavioural and biomarkers models. Panel a: ROC curves, with AUC values across models (AD vs HC). Panel b: Confusion matrices for the 4 models (AD vs HC). Panel c: ROC curves, with AUC values across models (nonAD vs HC). Panel d: Confusion matrices for the 3 models (nonAD vs HC). Panel e: ROC curves, with AUC values across models (AD vs nonAD). Panel f: Confusion matrices for the 3 models (AD vs nonAD). FN = false negatives, FP = false positives, TN = true negatives, TP = true positives.

In the non-AD versus HC comparison, the best behavioural model for predicting group (‘Behavioural’), consisted of a combination of TMT (β = 6.82, p < 0.001), Identification Accuracy (OMT) (β =-1.01, p = 0.032), and Localisation time (OMT) (β =-1.74, p = 0.063), with an AUC of 0.926 (CI 0.87-0.98), (**Figure 4**, panel c and d and Extended data). Localisation time was not included in subsequent analyses as not statistically significant in isolation. When biomarkers were used as predictors, the best ‘Biomarkers’ model consisted of a combination of PRDX6 (β =-1.02, p < 0.001) and NEFL (β = 0.85, p = 0.034), with an AUC of 0.886 (CI 0.82-0.95). In contrast, the ‘Combined’ model had an AUC of 0.968 (0.944-0.99), which, also in this cohort, was better compared to the ‘Behavioural’ and ‘Biomarkers’ models alone (χ² = 6.75, p-value = 0.009 and χ² = 14.3, p-value <0.001).

When comparing AD to non-AD patients, the best ‘Behavioural’ model consisted of Localisation time (OMT) (β = 1.07, p = 0.006), with an AUC of 0.760 (CI 0.63-0.89), with AD patients having longer reaction times compared to the non-AD group (**Figure 4**, panel e and f and Extended data). The best ‘Biomarkers’ model consisted of a combination of pTau231 (β = 3.36, p < 0.001), and ACHE (β = 1.94, p = 0.053) with an AUC of 0.905 (CI 0.89-1). Finally, the ‘Combined’ model had an AUC of 0.961 (CI 0.914-1). Unlike previous comparisons, in this cohort, the ‘Combined’ model was better compared to the ‘Behavioural’ model (χ² = 4, p-value = 0.045) but not compared to the ‘Biomarkers’ model (χ² = 2, p-value = 0.157).

### Network analysis

We further characterized the association between biomarkers through network analysis using the EBICglasso (Extended Bayesian Information Criterion Graphical LASSO) method. A graphical representation of the network can be found in **Figure 5**.

**Figure 5.**
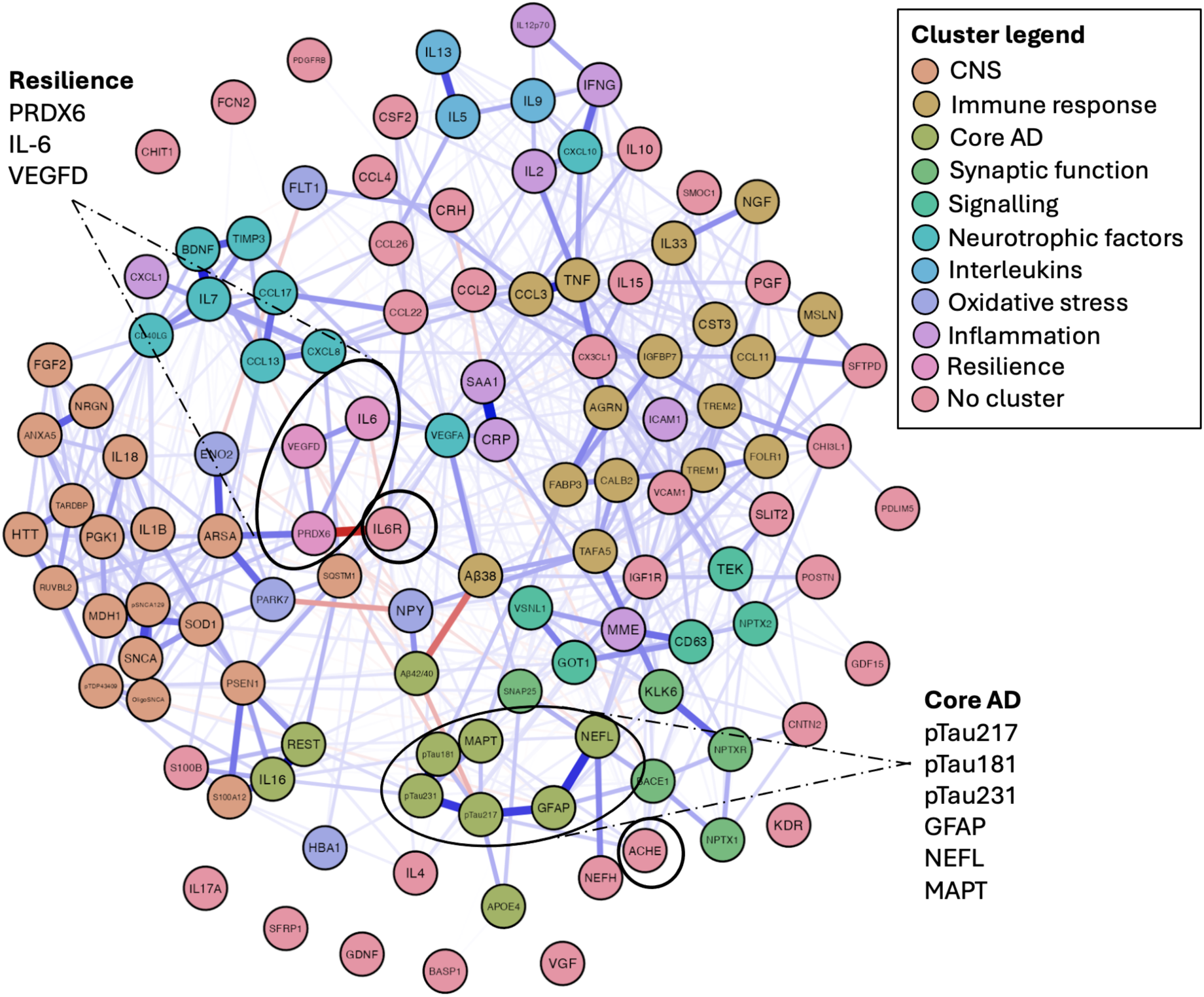
Network analysis: global plot. Positive weights between biomarkers are shown in blue, negative weights in red. The thickness of the weight represents the strength of the association. Different PC clusters are represented by different colours. The black circles highlight key proteins highlighted by this study.

Subsequently, we calculated *‘Betweenness’* centrality measures for each biomarker, which quantify how often a given node lies on the shortest paths connecting other nodes in the network. In other words, it reflects the extent to which a variable serves as intermediary between other variables. The 10 biomarkers with higher and lower *Betweenness* centrality are presented in Extended data. In brief, TNF was the biomarker which across all the network exhibited the highest *Betweenness* value, indicating its role as key mediator in the network. PRDX6, IL6R and pTau217 also showed high values. Conversely, ACHE had a very low Betweenness value, the lowest across all biomarkers, indicating its independence from most biomarkers in the network. This finding is in keeping with ACHE not being part of the first 10 PCA clusters and being in the periphery of **Figure 5**.

*Weights* between the three key biomarkers identified by the previous model comparison analysis (pTau217, PRDX6 and ACHE) and other biomarkers in the network were also computed by the network analysis and are presented in **Figure 6**. Positive weights indicate positive relationships, whereas negative weights indicate inverse correlations. pTau217 was strongly positively correlated with pTau231 and GFAP, moderately to ApoE4, pTau181, MAPT, ACHE, and to NEFL, CALB2 and interleukin 6 (IL6), while it was negatively correlated with PRDX6 and the Aβ42/40 ratio. This reinforces the role of PRDX6 as a protective factor. PRDX6 was positively associated with Arylsulfatase A (ARSA), IL6 and vascular endothelial growth factor D (VEGF-D), with the latter ones being part of the same PC cluster (*Resilience*), and the first being in the shared PC1 (CNS) PC. It also had a strong negative association with IL6R, and negative associations with pTau217, NEFL and MAPT. Conversely, ACHE was positively associated with pTau217, and to other proteins such as BACE1, MAPT and neuronal pentraxin 2 (NPTX2).

**Figure 6.**
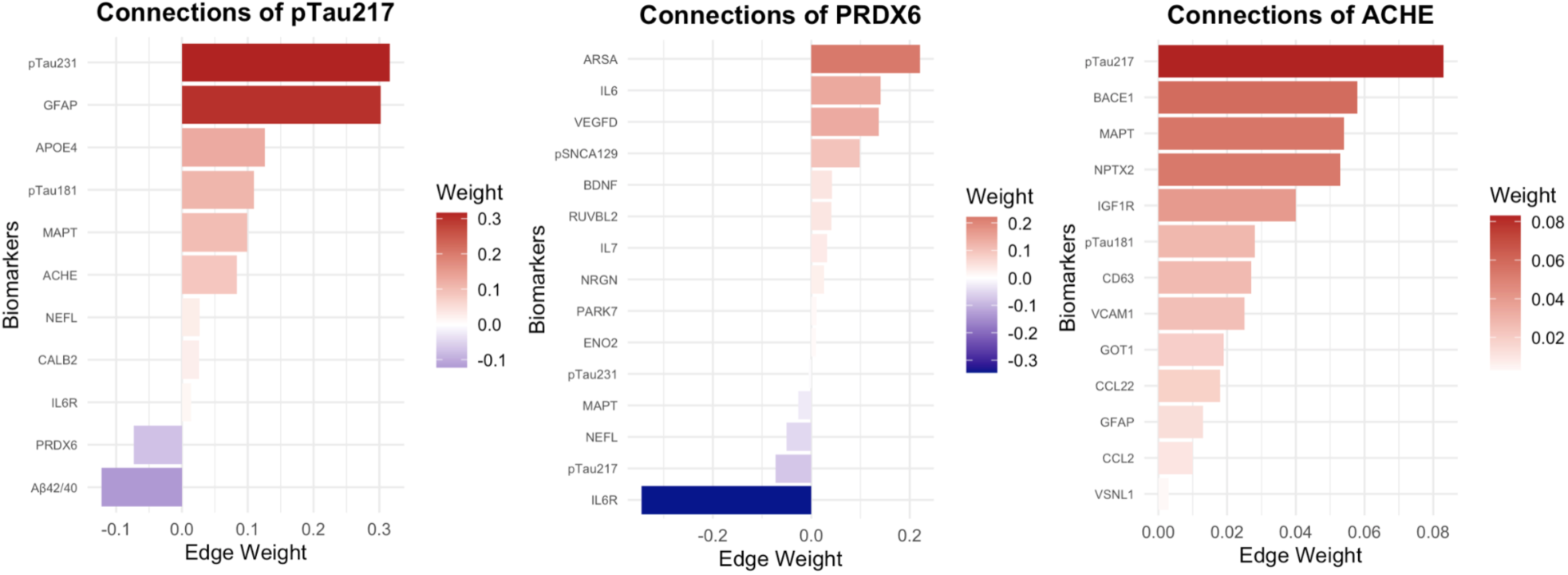
Network analysis: key biomarkers’ weights. Positive weights between biomarkers are shown in red, negative weights in violet. The darker the colour of the weight and the longer the bar, the stronger the association.

### Differential biomarkers’ profile between groups

Finally, we tested whether the key proteins found in previous analyses were exclusively higher/lower in patients with AD, or if they were over/underexpressed also in patients with other types of dementia. The comparison between AD and HC showed an upregulation in AD of multiple biomarkers, including but not limited to pTau species (pTau217, pTau231, pTau181), GFAP, NEFL, MAPT, ACHE and IL6R (**Figure 7**). Downregulated proteins in AD compared to HC included PRDX6, the Aβ42/40 ratio, VEGF-D, ARSA, IL6 and neuron specific enolase (ENO2). In patients with DLB, pTau231, and to a lesser extent pTau217 and IL6R were increased compared to HC.

**Figure 7.**
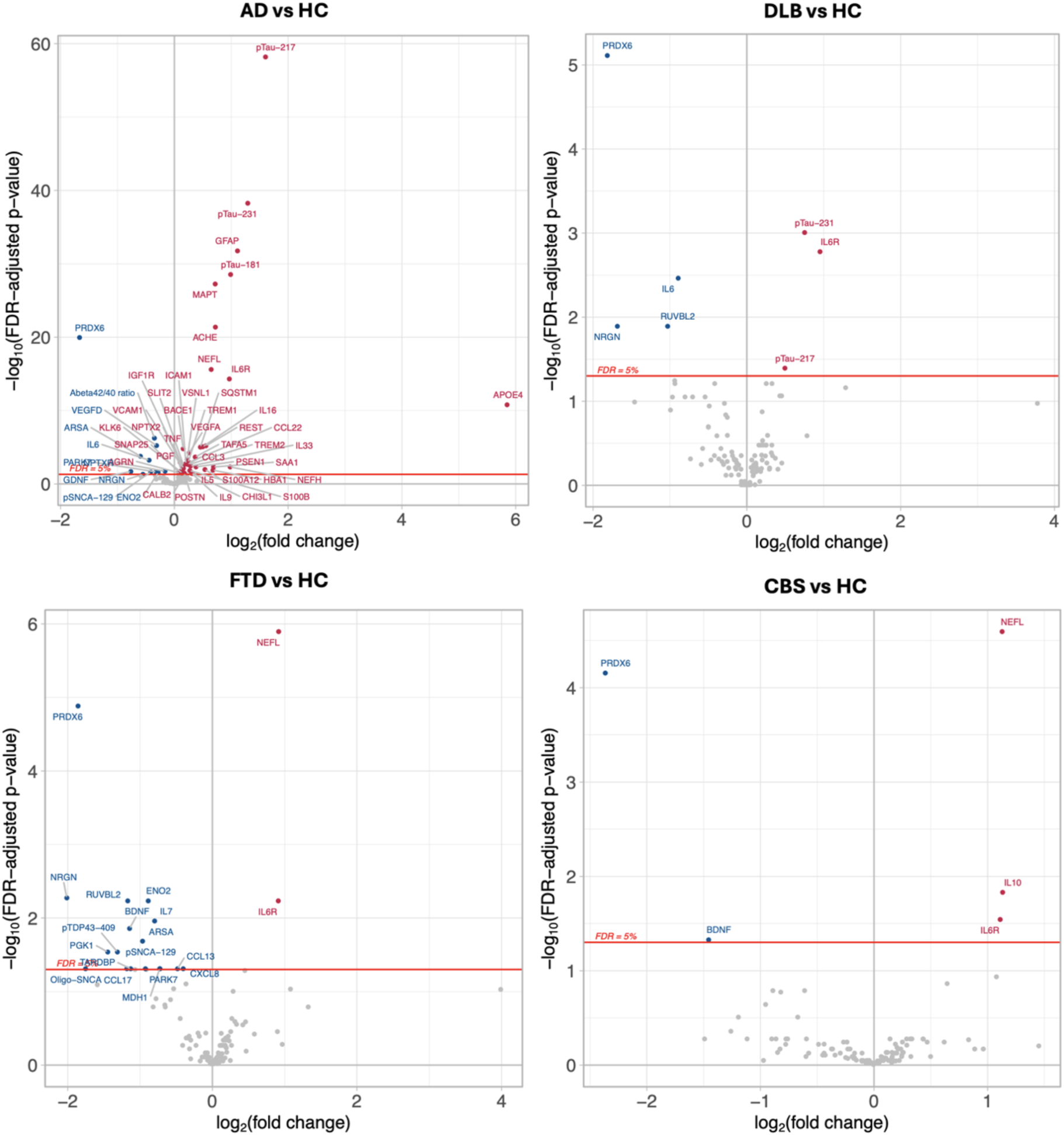
Plasma biomarkers levels in different groups (AD, DLB, FTD and CBS) versus healthy controls. Hypoexpressed proteins in blue, hyper in red. All results are corrected for multiple comparisons (False Discovery Rate-FDR).

**Figure 8.**
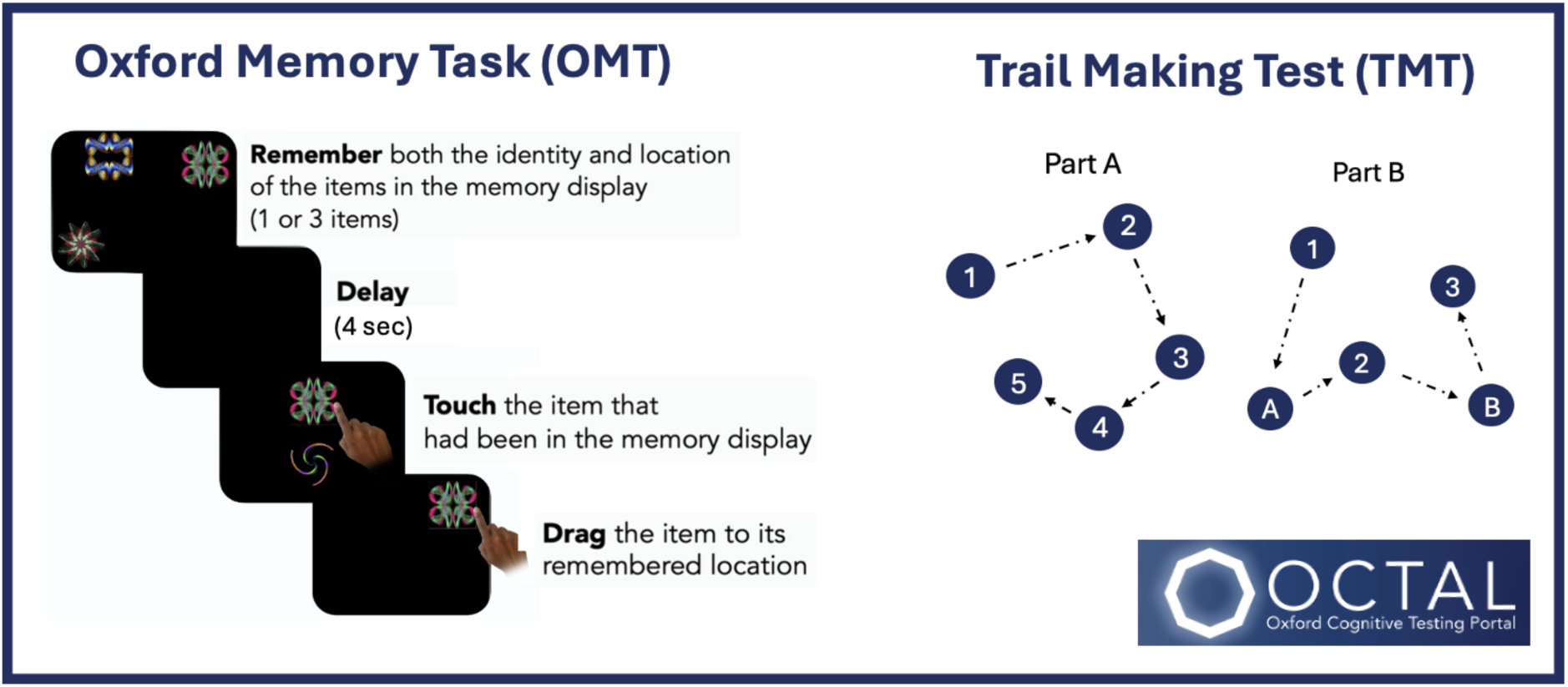
Oxford Cognitive Testing Portal (OCTAL) task design. Panel A: **Oxford Memory Test** task design. Participants were presented with either 1 or 3 fractals randomly distributed on the screen. After a 4 second delay two fractals appeared at the centre of the screen, one of which had appeared in the memory array whereas the other one was a distractor. Firstly, they needed to identify the object they had seen previously (‘what’) and then drag it back to its original location (‘where’). Several metrics can be extracted from this task, such as: **Identification accuracy**: the proportion of correctly identified items, **Absolute Error**: the distance between the original item and the participant’s response location, **Identification Time**: the time in seconds taken to identify the correct object, **Localization Time**: the time in seconds to drag the chosen object to its remembered location, **Misbinding**: the probability of mislocalizing a correctly identified item to the remembered location of another item in the memory array, **Guessing**: the probability of random guessing. **Imprecision**: the width of the distribution of the responses around the target. Panel B: A computerized version of the **Trail Making Test** is available on the platform. The average of part A and part B was used as the outcome metric.

Similarly to AD, PRDX6 was the protein which showed the highest effect size as greatly downregulated in DLB patients compared to healthy controls, and similarly IL6 was also reduced. However, in this case the Aβ42/40 ratio values were not different between DLB and HC groups. Other proteins such as neurogranin (NGRN) and RUVBL2 were hypoexpressed in DLB patients compared to healthy controls.

The comparison between FTD and CBS patients and healthy controls showed an increase in NEFL and IL6, but unlike AD and DLB, pTau species were not increased. PRDX6 was also in this case the protein which showed the lowest value in patients compared to healthy controls for both groups. BDNF was also reduced in both patient groups compared to controls, as well as several other proteins in FTD patients.

Similarly to the findings in neurodegenerative patients, the comparison between SCD and HC showed an overexpression of several different proteins including PRDX6, IL6, VEGF-D, ENO2, ARSA and IL6 in HC (see Extended data). PRDX6 and IL6 were also lower in AD compared to MCI patients, whilst several other biomarkers were increased, including core AD biomarkers (pTau217, pTau231, pTau181, GFAP, NEFL, MAPT), synaptic markers such as NPTX2, Sinaptosomal Associated Protein 25 (SNAP25) and other proteins including ACHE, BACE1, IGFR1R, SLIT2, VCAM1 and ApoE4.

When other neurodegenerative patient groups (DLB, FTD, CBS), were directly compared to the AD cohort, there was an increase in AD of the three pTau species (pTau217, pTau231, pTau181), of MAPT and ACHE (see Extended data). GFAP was raised in AD patients compared to DLB and FTD patients but not if compared to CBS patients and NEFL was not different across groups. CCL13 was raised in AD patients compared to FTD and CBS patients but not if compared to DLB. The Aβ42/40 ratio was lower only in AD patients compared to FTD.

## Discussion

Plasma biomarkers have the potential of changing current screening and diagnostic practice across neurodegenerative diseases, due to their non-invasive nature and low cost. In this study, we explored the relationship between a vast array of plasma biomarkers and cognition using a fully digitized cognitive test portal (OCTAL). Whilst several digital platforms have shown to be able to capture signs of cognitive impairment and relate to amyloid status, most data are limited to cognitively unimpaired individuals ^41,42^, or use CSF or PET benchmarks ^43^. The few studies available combining digital and plasma biomarkers are usually limited to a small number of biomarkers, such as pTau217 ^23^, and do not explore their performance in other forms of dementia. This is the first study, to our knowledge, using remote digital measures and a large panel of plasma biomarkers in a mixed memory clinic cohort.

We found that measures of localisation accuracy (Absolute Error, memory Imprecision and Localisation Time) showed the highest association to plasma biomarkers across standard linear correlations (**Figure 2**) and canonical correlations (**Figure 3**). We were also able to show that the combination of behavioural measures (Absolute Error (OMT) + Identification accuracy (OMT) + TMT) was as good as pTau217 in discriminating between HC and patients with AD (**Figure 4**). In that cohort, a model combining digital and plasma biomarkers achieved an AUC of 0.99 and was better compared to models with digital and plasma biomarkers if used in isolation. In the comparison between non-AD dementias and HC similar behavioural measures such as Identification accuracy (OMT) and TMT were important in clinical classification, whilst NEFL and PDRX6, and not pTau217, were the best performing biomarkers. In contrast, when trying to discriminate between patients with different forms of dementia (AD versus non-AD), measures such as Localisation time seemed to perform best. A combination of pTau231 and ACHE was selected by the model as the best performing combination in discriminating between different forms of dementia, likely because the model selection algorithm maximises the fit of the combination of different markers, rather than looking at markers in isolation.

Automated platforms offer the advantage of exploring multiple pathologic pathways simultaneously, in a quick and highly reproducible fashion, moving beyond single biomarkers’ quest. NULISA offers such a comprehensive proteomic approach, with its CNS panel measuring over 124 neurologically relevant proteins at once. However, a large array of biomarkers brings its own challenges. Dimensionality reduction through Principal component analysis allowed us to extract meaningful patterns within the digital and plasma biomarkers examined.

PC3 (*Core AD)*, showed the highest positive association with cognitive performance (**Figure 2**), where the higher the biomarkers’ values the worse the cognition across both tasks examined (OMT and TMT). Relationship between this cluster and behavioural measures were particularly strong for measures of spatial precision (i.e. Absolute Error, Imprecision and Localisation time). No significant correlation was found between Identification accuracy at the OMT and the Core AD cluster after correcting for multiple comparisons, further highlighting the importance of using precision measures that do not rely simply on a binary correct/incorrect answer ^44^.

PC9 (*Inflammation*), was also strongly associated with cognitive performance, across *Spatial precision*, *Spatial identity*, *Executive functions*, but not *Reaction times* PCs, and was also negatively associated with *Accuracy*, where the higher the inflammatory biomarkers levels, the worse the accuracy at the OMT task. Pro-inflammatory markers such as IFN-γ and C-reactive protein (CRP) have been previously reported as increased in different forms of dementia ^45^, and to be associated with worse cognitive performance, particularly at tests measuring executive function, though these are not specific to AD ^46,47^. The data presented here support a detrimental impact of these pro-inflammatory markers on cognitive function in a mixed memory clinic cohort. Lastly, PC5 (*Signalling*) showed a positive correlation with the *Spatial precision* PC only.

Conversely, PC1 (*CNS)*, PC4 (*Synaptic function)* and PC10 (*Resilience)* showed negative correlations with cognitive performance, where the higher the values, the better the cognitive performance. PC1 (*CNS)*, despite being the largest cluster of plasma biomarkers, was not the one showing the largest correlations with cognitive performance (**Figure 1 and 2**). ARSA, NRGN, RUVBL2, and PRDX6 were part of that cluster and were also overexpressed in HC compared to other patient groups (**Figures 7** and Extended data), pointing towards a protective role of these proteins.

PRDX6 emerged as a key protein for cognitive resilience, as the only protein with negative loading in the canonical correlation analysis (**Figure 3**). PRDX6 was part of both PC1 and PC10 (**Figure 1**), which could explain the negative correlation between CNS cluster values and cognitive performance (**Figure 2**). It was also selected as amongst the three best performing biomarkers, together with pTau217 and ACHE in discriminating between HC and AD (**Figure 4**) and also emerged as key biomarker in discriminating between HC and non-AD dementias. Unlike pTau217 and ACHE, it had a negative β coefficient, suggesting a protective effect. This was also shown by the negative weights in the network analysis (**Figures 5 and 6**) and overexpression of PRDX6 in HC compared to all patient groups (**Figure 7** and Extended data). PRDX6 is a protein with antioxidant effects, involved in the repair of oxidatively damaged cell membrane lipids and cellular signalling ^48^. It is expressed in the CNS exclusively by astrocytes. Overexpression of PRDX6 in APPswe/PS1dE9 AD transgenic mice has been found to cause suppression of plaque seeding, remodeling of mature plaques, reduced brain Aβ load and Aβ-associated neuritic degeneration^48^. It is however the first time it has been described as a protective factor for cognitive impairment in humans.

Arylsulfatase A (ARSA), a lysosomal enzyme, is known to inhibit the aggregation and propagation of α-synuclein, and lower levels have been linked to more severe cognitive impairment in patients with Parkison’s Disease ^49^. Here, ARSA was the protein more closely related to PRDX6 at the network analysis (**Figure 6**), further suggesting a beneficial effect on cognition. Although a negative effect of high NRGN levels is often reported in CSF ^50, 51, 52^, evidence using NULISA showed reduced levels of NGRN in amyloid PET positive individuals compared to amyloid negative individuals ^53^. Our study supports a protective effect of NGRN on cognitive function, given its overexpression in cognitively healthy controls (**Figure 7** and Extended data) and positive correlation with PRDX6 (**Figure 6**). RUVBL2 levels in plasma have been previously associated with lower accumulation of tau-PET burden over time in cognitively healthy individuals ^54^. Our findings support this positive effect given the negative association with cognitive impairment and overexpression in HC (**Figure 7** and Extended data).

Cluster PC10 (*Resilience),* consisting of PRDX6, IL6 and VEGF-D, showed a protective effect on cognitive function, where the higher the level of the proteins, the better the performance (**Figure 2**). IL6 was higher in HC compared to the SCD, AD and DLB groups, whilst VEGF-D was increased in SCD and AD compared to HC (**Figure 7** and Extended data). Conversely, IL6R was overexpressed in all patient groups compared to HC. IL6R was also in the top biomarkers related to cognitive performance at the canonical analysis (**Figure 3**), with the higher the levels the worse the cognition. IL6 and IL6R show key roles in the network analysis (**Figure 5**) and are respectively highly positively and negatively correlated with PRDX6 values (**Figure 6**). In previous studies, IL6 has been found to exert both positive and negative effects on neuron survival and cognition^55,56^. In our study levels of IL6 were lower whilst IL6R were higher in patients’ populations, suggesting a neuroprotective mechanism. High IL6R levels have been reported in AD and other neurodegenerative diseases in a large proteomic study^57^. Our study supports these transdiagnostic findings. VEGF-D contributes to maintenance of dendritic arborization and has been studied for its neuro-protective effects against neuronal damage in stroke ^58, 59^. It has also been shown to promote hippocampal neurogenesis and improve learning and memory performance in a mouse model ^60^. Our findings support its protective role as hyperexpressed in HC and linked to PRDX6 (**Figure 6 and 7**).

ACHE levels were also found to be informative in discriminating AD dementia patients compared to other groups (**Figure 7** and Extended data). They also showed a high impact on cognition at the canonical correlation analysis (**Figure 3**) and were amongst the 3 top biomarkers together with pTau217 and PRDX6 in group classification between HC and AD and non-AD dementias (**Figure 4**). This is in line with recent findings using SomaScan, which showed ACHE as a key upregulated protein in AD with high effect size^10^. We also showed that whilst closely linked to pTau217 levels (**Figure 6**), it had a very low betweenness centrality value (**Figure 5** and Extended data), i.e. was not closely correlated to most other biomarkers. The reasons for such a unique profile need to be unravelled by future studies.

The analyses of group differences showed common comparable top upregulated proteins in AD to several studies using NULISA technology and other platforms, such as pTau217, pTau231, pTau181, GFAP, NEFL and MAPT ^53,15^. However, some proteins such as IL6R and ACHE have been less frequently reported using single assays, but can be unravelled by proteomics panels using NULISA or SomaScan technology^10^. The increased levels of NEFL in FTD have been reported before using different platforms, including NULISA ^15,57^, but such findings in CBS patients have not been previously described. We were also able to replicate the findings that in patients with DLB, pTau231, and to a much lesser extent pTau217 is increased compared to HC ^15^.

This study has some limitations; the sample size of the patient cohorts is small, and replication of these findings on a larger dataset is warranted. Further, the behavioural metrics used in this study had a significantly lower performance in discriminating between AD and non-AD dementias (**Figure 4**). This might be expected given the cognitive tests used in this study tap into cognitive processes, e.g., short-term memory and executive functions, which are impaired across different neurodegenerative diseases and are not specific to AD. Another key consideration is that most AD and non-AD dementia patients were able to have a blood sample taken, whilst not all completed cognitive testing (see Methods), largely due to their degree of cognitive impairment. This was in contrast with healthy controls, who had much higher completion rates. A higher sample size is needed in order to unravel which combination of digital and plasma biomarkers metrics performs best in discriminating between different forms of dementias. Expanding the range of cognitive processes studied across different neurodegenerative diseases would also be key to find a ‘digital’ behavioural signature of cognitive impairment, analogous to their pen-and-paper counterparts.

Plasma biomarkers obtained using NULISA’s CNS panel provide key insights into several biological pathways across different types of dementia in an easy and automated fashion and can relate to digital markers of cognitive impairment. Key proteins related to cognitive functions (both detrimental and protective) can be studied with such platforms. No single biomarker or cognitive measure will ever substitute clinical assessment in diagnosing a neurodegenerative disease. However, we strongly advocate the integration of both digital measures and plasma biomarkers in existing care pathways to assist clinical decision-making.

## Online Methods

### Participants

Three hundred ninety one participants took part in the study; Alzheimer’s disease dementia (AD) (n=88), Mild cognitive impairment (MCI) (n = 28), Subjective cognitive decline (SCD) (n=31), Lewy body disease (DLB) (n=18), Corticobasal syndrome (CBS) (n = 9), Frontotemporal dementia (FTD) (n=16), and elderly healthy controls (HC) (n=201), (see Extended data for demographics’ and test scores).

HC, SCD, MCI, and AD patients were recruited across the Cognitive Disorders Clinic at the John Radcliffe Hospital in Oxford (Neurology cohort), Oxford Health NHS Foundation Trust (Warneford Hospital, Oxford and Whiteleaf Centre, Aylesbury) and Sussex Partnership NHS Foundation Trust as part of the Feasibility and Acceptability of Scalable Tests (FAST) Brain Health Study (FAST cohort). DLB, CBS and FTD patients were recruited from the Cognitive Disorders Clinic at the John Radcliffe Hospital in Oxford. Participants underwent blood collection in person, online remote digital cognitive testing, and face-to-face standard cognitive testing (the latter in the Neurology cohort only).

For the Neurology cohort, SCD was defined according to the 2020 criteria from Jessen et al. for subjective cognitive decline^34^. MCI patients were classified according to Petersen’s criteria of 2014^35^. AD dementia patients were defined as having AD clinical syndrome according to the 2018 ATN criteria^1^ and will be subsequently referred to as AD. They had a progressive, multidomain, cognitive impairment and underwent MRI brain imaging, FDG-PET imaging and CSF if required, the results of which were in keeping with a clinical diagnosis of AD. Patients with FTD, DLB and CBS were diagnosed according to Rascovsky’s criteria^36^ (FTD), McKeith criteria (DLB)^37^ and Armstrong’s criteria (CBS)^38^. Elderly healthy controls were >50 years old, had no psychiatric or neurological illness, were not on regular psychoactive drugs, and all scored above the cut-off for normality (88/100 total Addenbrooke’s Cognitive Examination-III (ACE-III) score). For the FAST cohort, diagnostic labelling was based on self-report according to study protocol.

### Ethics

The study was performed in accordance with the ethical standards as laid down in the 1964 Declaration of Helsinki and its later amendments. Ethical approval was granted by the University of Oxford ethics committee (IRAS ID: 248379, Ethics Approval Reference: 18/SC/0448 and IRAS ID: 301319, Ethics Approval Reference: 22/WA/0183). All participants gave written informed consent prior to the start of the study.

### Blood sample collection and processing

Blood was collected in ethylenediaminetetraacetic acid (EDTA) tubes (10 mL each), and centrifuged within 30 minutes (1800 g, room temperature, 10 minutes). The EDTA tubes were gently inverted after collection to avoid coagulation. After centrifugation, plasma was aliquoted into 0.5 mL polypropylene tubes (Fluid X, Tri-coded Tube, Azenta Life Sciences) and transferred into a −80◦C freezer. All cryovials were anonymized, and the unique cryovial code was logged into a secure database, linked to the participant’s anonymous code and visit number. Samples were shipped in dry ice to Alamar Biosciences, Fremont, CA. Sample and data analysis was performed according to kit manufacturer protocols, including Log2 Transformation of the data and NULISA Protein Quantification (NPQ) on the logarithmic scale provided by Alamar Biosciences.

### Digital cognitive assessment

Participants completed two digital cognitive assessments from OCTAL (Oxford Cognitive Testing Portal, available at https://octalportal.com), the Oxford Memory Test and Trail Making Test. The platform and details of cognitive tests have been described in detail elsewhere ^24,25,29^, and are briefly summarized below (Figure 1).

### Oxford Memory Test

Oxford Memory Test (OMT) is a short-term memory task, which has been deployed across several neurodegenerative diseases ^26,27,28,29,30,31,32,33^. Participants were presented with one or three fractal patterns positioned at various locations on screen for 3 seconds. After a 4 second delay one of these fractal patterns was shown alongside a foil pattern. Participants were asked to identify the fractal they saw and drag it back to its original location.

Overall, this task allowed to extract seven different working memory metrics, including:

- **Identification Accuracy**: the proportion of trials in which participants correctly identified the previously presented item.
- **Absolute Error**: the distance from original item location to participant’s response location.
- **Identification Time**: the time in seconds taken to identify the correct object.
- **Localization Time**: the time in seconds to drag the chosen object to its remembered location.
- **Misbinding**: the probability of mislocalizing a correctly identified item to the remembered location of another item in the memory array.
- **Guessing**: the probability of random guessing.
- **Imprecision**: the width of the distribution of the responses around the target.

### Trail Making Test

The Trail Making Test (TMT) measures processing speed and executive functions. In this online version, 25 circled numbers were presented on screen, and participants were instructed to connect them by clicking the circles in order as fast as possible. It consisted of three trials of Task A (where the order is 1-2-3-4-5-6-…) and three trials of Task B (order 1-A-2-B-3-C-…). The average of version A and B was used for analyses.

As participants did the task remotely with their own devices, the size of stimuli was harmonized across different devices using a previously described card calibration procedure^39^. The platform is designed and managed by S.Z. The tasks are built using the PsychoPy Builder (PsychoJS, version 2022.2.4) with custom-written codes in Javascript and hosted on the server system Pavlovia.org. A link with a unique patient and visit identifier was sent to the participants’ e-mail address the same day as the in-person visit when blood was collected. Participants could use a desktop, laptop, or tablet with any operating system to complete the tests.

## Statistical analysis

### Demographics and cognitive tests

For analysis and data visualization purposes R studio (version 4.4.1), Python (3.7.6) and JASP (version 0.19.3), were used. Demographics and cognitive tests were compared using Kruskal-Wallis tests, with Dunn’s post-hoc tests. P-values were two-tailed with statistical significance set at p < 0.05 for all analyses. NULISA Protein Quantification (NPQ) plasma biomarkers’ values represent already Log2 transformed values, whilst digital cognitive measures were z-scored prior to analysis.

### Principal component analysis

Firstly, a principal component analysis was performed on both digital behavioural measures and plasma biomarkers. Oblique promax rotation was used, with the number of components (PC) based on parallel analysis, and base decomposition calculated on their correlation matrix.

The scree plot of the biomarkers’ PCs can be found in Supplementary materials. For biomarkers, only factor loadings > 0.4 were retained.

Secondly, linear correlations between PC of digital and plasma biomarkers were evaluated. These were assessed with Spearman rank test, using age, sex, and education as covariates. The Benjamini–Hochberg method, which controls the false discovery rate (FDR), was used to correct for multiple comparisons.

### Canonical correlation analysis

Thirdly, a canonical correlation analysis (CCA) between digital and plasma biomarkers was computed. CCA is a multivariate statistical technique that explores the relationships between two groups of variables. The method constructs pairs of canonical components, each being a weighted linear combination of the variables within one set. These are derived so that the correlation between the two canonical components - one from each set—is maximized. The first pair of components captures the strongest shared association between the two sets; subsequent pairs capture remaining associations orthogonal to previous ones.

### Logistic regression

Subsequently, we applied logistic regression using the R-based MuMIn (multi-model inference) package to test which combinations of behavioural measures and biomarkers would best predict group classification between 1) healthy controls and patients with AD, 2) healthy controls and patients with non-AD dementias (DLB, FTD and CBS combined) and 3) AD and non-AD dementias. Subgroup comparisons were not tested given the low numbers in each group with complete (behavioural and biomarkers) datasets. For biomarkers, only factors with a loading of > 0.6 on the canonical correlation were retained. The *dredge* function provided in the MuMIn package was used to achieve model selection, with the best-performing model having the lowest corrected Akaike information criterion (AIC). This was run separately for behavioural measures and plasma biomarkers. Model performance was evaluated using AUC - the area under the receiver operating characteristic curve (ROC), sensitivity, specificity, positive predictive value (PPV), negative predictive value (NPV), and accuracy. Predicted probabilities were obtained from the logistic regression model and evaluated against the observed binary outcome (belonging to the assigned group). Because the groups were imbalanced, the optimal probability threshold for classification was determined using Youden’s J statistic. AUC values are presented with their corresponding 95% confidence interval (CI). Beta (β)-coefficients are reported for each parameter in composite models. Pairwise McNemar χ² test was used to compare accuracy between models.

### Network analysis

Additionally, we performed a network analysis using the EBICglasso (Extended Bayesian Information Criterion Graphical LASSO) method as implemented in JASP (JASP Team, 2025). A weighted network was estimated, where each *node* represented a biomarker and edges (connections) reflected the regularized partial correlations between pairs of biomarkers, computed using the graphical least absolute shrinkage and selection operator (EBICglasso) method. It builds on the LASSO (Least Absolute Shrinkage and Selection Operator) method, which introduces a penalty on the precision matrix (the inverse covariance matrix). It shrinks small off-diagonal elements toward zero, and if they are close enough to zero, sets them exactly to zero. This produces a sparse network, which is one that contains only the most robust conditional relationships. *Weights* quantify the unique association strength between two biomarkers after accounting for all other variables in the network. Stronger absolute weights indicate a more robust relationship. To examine the relative importance of nodes, we computed *betweenness* as a centrality index, which captures the extent to which a given biomarker lies on the shortest paths connecting other biomarkers. Nodes with higher betweenness centrality can be interpreted as key intermediaries that facilitate the flow of associations or interactions within the network.

Finally, plasma biomarkers patterns across diagnostic groups were compared using a linear mixed effect model, with age and gender as covariates.

### Quality control analysis

The average median intra-plate and inter-plate assay coefficient of variation (CV) were respectively 4.65% and 6.46%, well within recommended expected ranges (below 10% and 15%). NULISA markers were detected with high sensitivity (95.2%). 7 samples did not pass Alamar’s quality control, i.e., very high deviation (>95%) of the internal control median, and were excluded from further analyses. 5 out of 124 proteins (GDI1, PTN, SNCB, UCHL1, YWHAZ) did not meet detectability criteria and were therefore discarded. To aid interpretability, the Aβ42/40 ratio was calculated from the NPQ values of Aβ42 and Aβ40 according to the manufacturer’s instructions and individual values of Aβ42 and Aβ40 were not included in further analyses. A total of 375 subjects had available biomarkers data and 299 completed digital cognitive assessment; Alzheimer’s disease dementia (AD) (n=43), Mild cognitive impairment (MCI) (n = 23), Subjective cognitive decline (SCD) (n=28), Lewy body disease (DLB) (n=8), Corticobasal syndrome (CBS) (n = 6), Frontotemporal dementia (FTD) (n=9), and elderly healthy controls (HC) (n=182).

## Data Availability

Data are available upon reasonable request. Access is contingent upon agreement with the original data providers (i.e. the researchers responsible for data collection) and subject to applicable ethical, legal, and confidentiality constraints.

## Acknowledgements

This work was supported by the Wellcome Trust and the National Institute for Health Research (NIHR) Oxford Health Biomedical Research Centre. S.Z., S.T., M.B., A.S., A.F., M.H. are funded by the Wellcome Trust [226645/Z/22/Z]. S.T., S.G.M. and M.H. are also funded by the NIHR Oxford Health BRC. The project was further supported by a Guarantors of Brain post-doctoral fellowship to S.T. S.G.M. is supported by the NIHR Oxford BRC. C.G. received a residency scholarship, funded by the Italian Ministry of University and Research (MUR), as part of the Neurology Residency Program at the Università degli Studi di Milano. A.F. is further supported by funds from Medical Research Council [MR/X022013/1] and MyAware. CMvD is supported by the US National Insititute on Aging (NIH), NovoNordisk, the Oxford-GSK Institute of Molecular and Computational Medicine (IMCM), Centre of Artificial Intelligence for Precision Medicines (CAIPM) of the University of Oxford and King Abdul Aziz University, Alzheimer Research UK (ARUK), UK National Institute for Health and Care Research (NIHR) Oxford Biomedical Research Center (BRC), ZonMW (Delta Dementie) and Alzheimer Nederland. She is currently the Research Director Brain Health of the Health Data Research UK (HDR UK) and the UK Dementia Research Institute (UK DRI), working in partnership with Dementias Platform UK (DPUK). I.K. declares funding for this work through Alzheimer’s Research UK, Alzheimer’s Society and the People’s Postcode Lottery READ OUT grant as well as the Medical Research Council (Dementias Platform UK) and the Oxford Health NIHR Biomedical Research Facility. The views expressed are those of the author(s) and not necessarily those of the funders.

