## Extended data for "Novel markers of cognitive impairment and resilience using digital and plasma biomarkers across different neurodegenerative diseases"

### Demographics

A summary of participants' demographics and test scores can be found below.

| | HC (n = 201) | SCD (n = 31) | MCI (n = 28) | AD (n = 88) | DLB (n = 18) | CBS (n = 9) | FTD (n = 16) | p-value |
|---|---|---|---|---|---|---|---|---|
| **Age** | 68.3 (8.4) | 63.0 (8.6) | 73.5 (8.6) | 69.7 (9.2) | 68.9 (7.1) | 72.3 (7.3) | 62.1 (9.7) | <0.001 |
| **Gender** | 78/123 | 12/14 | 16/12 | 40/48 | 15/3 | 5/4 | 12/4 | <0.001 |
| **ACE-III*** | 97.5 (1.9) | 95.2 (6.0) | 86.4 (5.9) | 59.9 (21.1) | 75.1 (12.7) | 81.6 (10.5) | 72.8 (12.2) | <0.001 |
| **IA (OMT)** | 0.90 (0.05) | 0.89 (0.05) | 0.82 (0.10) | 0.79 (0.10) | 0.74 (0.16) | 0.85 (0.07) | 0.82 (0.09) | <0.001 |
| **AE (OMT)** | 77.6 (40.5) | 78.7 (22.1) | 114.8 (46.5) | 185.8 (90.7) | 159.3 (58.1) | 127.9 (33.1) | 135.0 (130.6) | <0.001 |
| **MB (OMT)** | 0.28 (0.09) | 0.29 (0.08) | 0.35 (0.09) | 0.37 (0.10) | 0.42 (0.11) | 0.38 (0.04) | 0.37 (0.07) | <0.001 |
| **GS (OMT)** | 0.15 (0.08) | 0.16 (0.07) | 0.23 (0.09) | 0.31 (0.11) | 0.33 (0.12) | 0.33 (0.14) | 0.24 (0.11) | <0.001 |
| **Imp (OMT)** | 50.3 (28.7) | 53.2 (13.9) | 74.6 (26.7) | 119.8 (60.8) | 105.3 (45.8) | 94.1 (39.9) | 87.2 (91.3) | <0.001 |
| **IT (OMT)** | 2.56 (0.9) | 2.45 (0.6) | 3.0 (1.3) | 4.2 (2.1) | 3.7 (2.3) | 3.8 (1.2) | 2.5 (0.7) | <0.001 |
| **LT (OMT)** | 3.55 (1.7) | 2.66 (0.7) | 3.7 (1.3) | 6.4 (3.5) | 3.6 (1.4) | 2.7 (2.8) | 3.8 (1.5) | <0.001 |
| **TMT** | 34.4 (10.3) | 32.3 (8.6) | 58.9 (25.2) | 103.7 (101.7) | 156.7 (87.5) | 70.1 (28.9) | 59.1 (20.2) | <0.001 |

**Table 1S | Demographics and digital test results**
M = male, f = female. ACE-III = Addenbrooke's Cognitive Examination III. IA = identification accuracy, AE = absolute error, MB = misbinding, GS = guessing, Imp = imprecision, IT = identification time, LT = localisation time, TMT = trail making test. * data available for the Neurology cohort only

### Principal component analysis: plasma biomarkers

The first component, **PC1** (which we label as ***Central nervous system, CNS***), included several proteins involved across different neurodegenerative diseases, which are not specific to AD. The second component, **PC2** (***Immune response***), comprised several chemokines, cytokines and other proteins involved in immune signalling in the brain, which have been found to be increased in AD and other neurodegenerative diseases. The third component, **PC3** (***Core AD)***, included as positive loadings pTau species (pTau217, pTau231, pTau181), MAPT, GFAP, NfL, IL-16, RE1-Silencing Transcription factor (REST), ApoE4 and as negative loading the Aβ42/40 ratio. The fourth component, **PC4** (***Synaptic function***), encompassed proteins traditionally involved in synaptic function such as neuronal pentraxin 1 (NPTX1), neuronal pentraxin receptor (NPTXR) and SNAP25, as well as BACE1, which is crucial for Aβ cleavage and synaptic vesicle release, and Kallikrein 6 (KLK6). The fifth component, **PC5** (***Signalling***), included proteins involved in protein degradation such as membrane metallo-endopeptidase (MME), Glutamic-oxaloacetic transaminase 1 (GOT1) and calcium-sensor protein Visinin-like protein 1 (VSNL1). The sixth component, **PC6** (***Neurotrophic factors)***, encompassed different proteins such as Brain-derived neurotrophic factor (BDNF) and TIMP Metallopeptidase Inhibitor 3 (TIMP3), which play a major role in the survival of neurons and preservation of the extracellular matrix. The seventh component, **PC7** (***Interleukins)*** included several interleukins, which have shown to exert a protective role in AD. The eight component, **PC8** (***Oxidative stress***) consisted of different proteins involved in response to oxidative stress across different neurodegenerative diseases. The ninth component, **PC9** (***Inflammation***) comprised of several proteins involved in neuroinflammation, such as interferon gamma (IFN) and C reactive protein (CRP). Finally, the tenth component, **PC10** (***Resilience***) included PRDX6 (Peroxiredoxin-6), IL-6 and VEGF-D, proteins involved in oxidative stress, inflammation and vascular health.

### Principal component analysis: behaviour

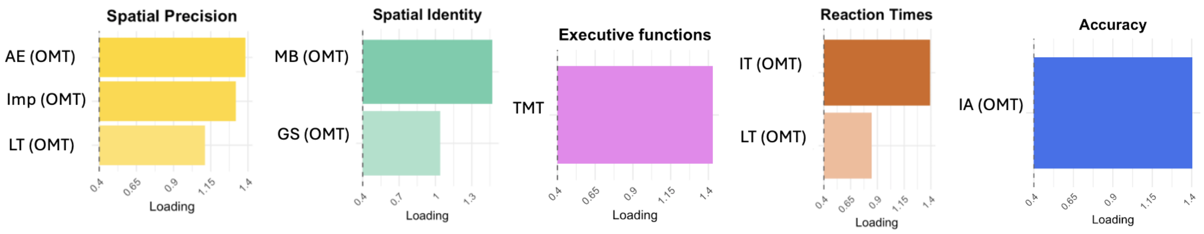


**Figure 1S | Principal component analysis: Factor Loadings for behaviour**
IA = identification accuracy, AE = absolute error, MB = misbinding, GS = guessing, Imp = imprecision, IT = identification time, LT = localisation time, TMT = trail making test.

| Cohort | Model | AIC | AUC (CI) | Accuracy | PPV | NPV | Sensitivity | Specificity |
|---|---|---|---|---|---|---|---|---|
| **AD/HC** | Behavioural | 93.9 | 0.933 (0.88-0.98) | 0.865 | 0.579 | 0.968 | 0.868 | 0.864 |
| | Biomarkers | 79.3 | 0.966 (0.93-1) | 0.921 | 0.706 | 0.988 | 0.947 | 0.915 |
| | Combined | 52.65 | 0.990 (0.98-1) | 0.972 | 0.9 | 0.989 | 0.947 | 0.977 |
| | pTau217 alone | 106.3 | 0.94 (0.90-0.98) | 0.907 | 0.688 | 0.97 | 0.868 | 0.915 |
| **nonAD/HC** | Behavioural | 69.5 | 0.926 (0.87-0.98) | 0.74 | 0.265 | 0.992 | 0.947 | 0.717 |
| | Biomarkers | 92.8 | 0.886 (0.82-0.95) | 0.714 | 0.247 | 0.992 | 0.947 | 0.689 |
| | Combined | 57.8 | 0.968 (0.944-0.99) | 0.841 | 0.38 | 1 | 1 | 0.825 |
| **AD/nonAD** | Behavioural | 64.4 | 0.760 (0.63-0.89) | 0.719 | 0.867 | 0.556 | 0.684 | 0.789 |
| | Biomarkers | 37 | 0.950 (0.89-1) | 0.93 | 0.947 | 0.895 | 0.947 | 0.894 |
| | Combined | 35.4 | 0.961 (0.914-1) | 0.93 | 0.925 | 0.941 | 0.974 | 0.842 |

**Table 2S | Model comparisons**
AIC = akaike information criterion, AUC = area under the curve, CI = confidence interval, PPV = positive predictive value, NPV = negative predictive value.

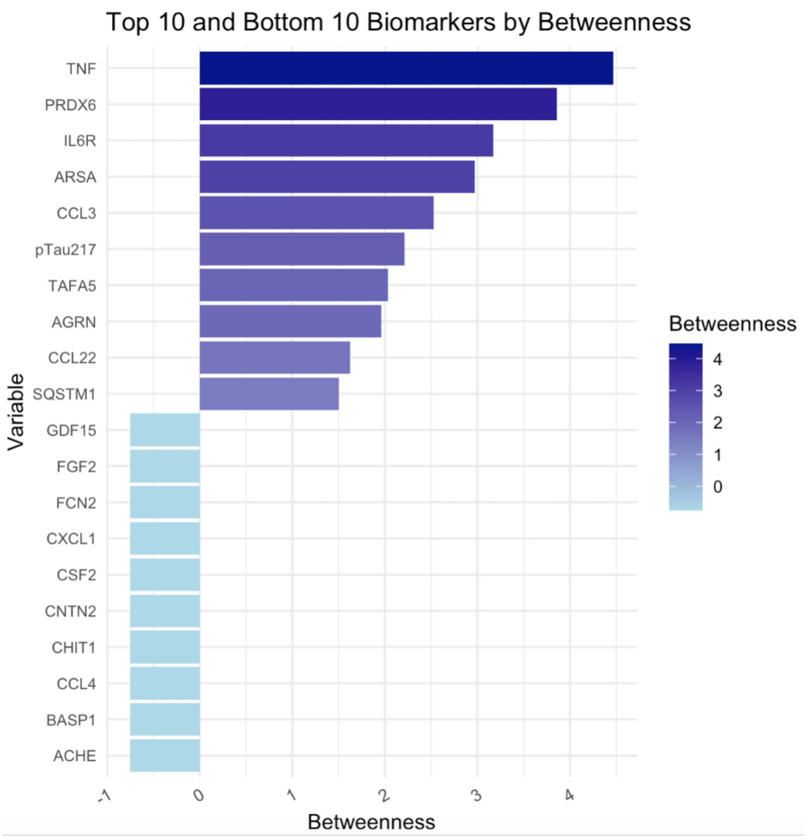


**Figure 2S | Network analysis: Betweenness**
Stronger loadings are represented by darker colours and longer bars.

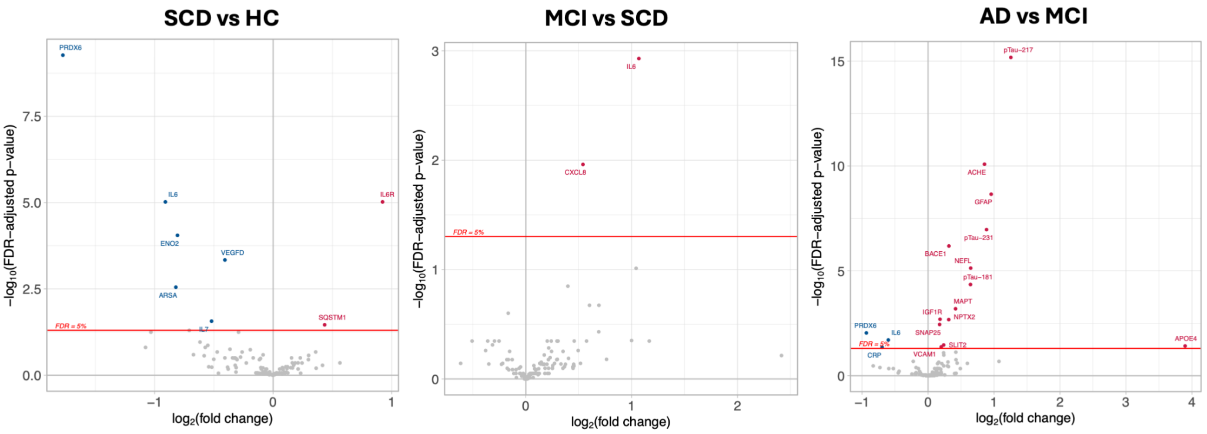


**Figure 3S | Plasma biomarkers levels across the AD continuum**
Hypoexpressed proteins in blue, hyper in red. All results are corrected for multiple comparisons (False Discovery Rate-FDR).

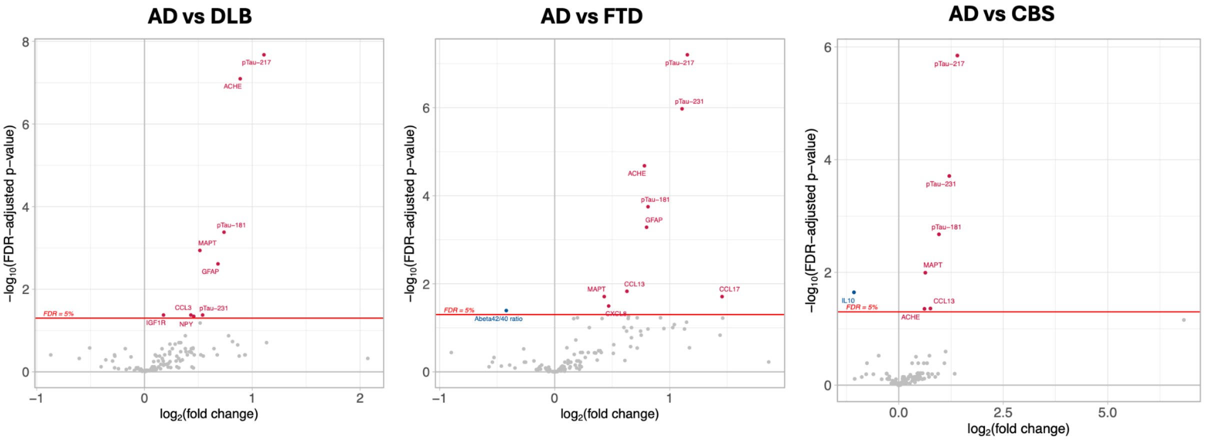


**Figure 4S | Plasma biomarkers levels across different dementia types (DLB, FTD and CBS) compared to AD**

Hypoexpressed proteins in blue, hyper in red. All results are corrected for multiple comparisons (False Discovery Rate-FDR).

### Digital profiling between groups

OCTAL's metrics captured different stages of the disease as well as discriminated between different forms of dementia (Figure 1S). Identification accuracy, Absolute Error, Guessing, and Misbinding on OMT were lower in patients with MCI as well as other primary dementias compared to controls. Trail making test also was lower in MCI and all the other dementias compared to healthy controls. Localization time was significantly higher only in patients with AD but not in other forms of dementia. Absolute Error, Identification Time and Localisation Time could also distinguish between MCI and AD dementia.

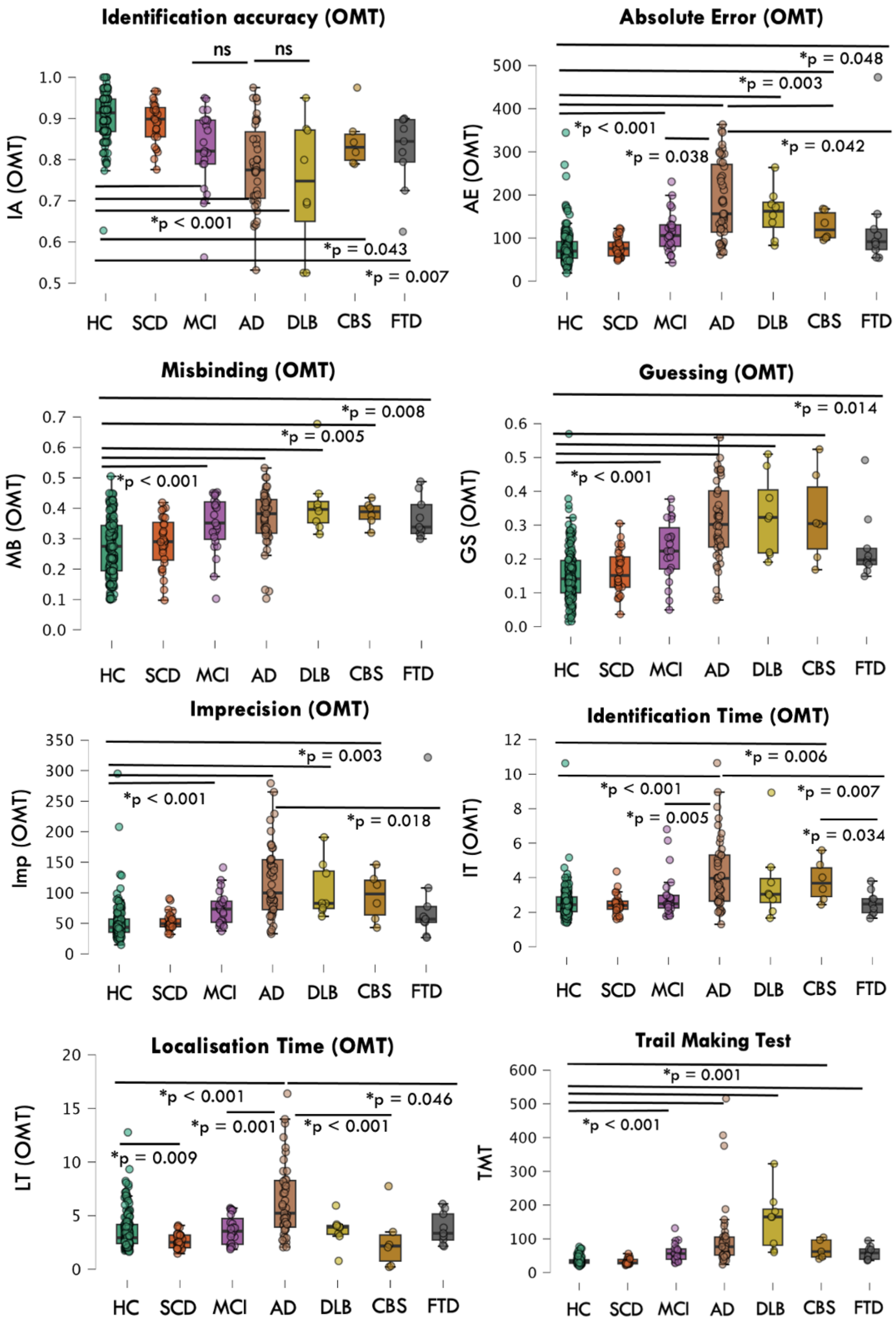


**Figure 4S | Digital Cognitive assessment across different neurodegenerative diseases**
IA = identification accuracy, AE = absolute error, MB = misbinding, GS = guessing, Imp = imprecision, IT = identification time, LT = localisation time, TMT = trail making test.

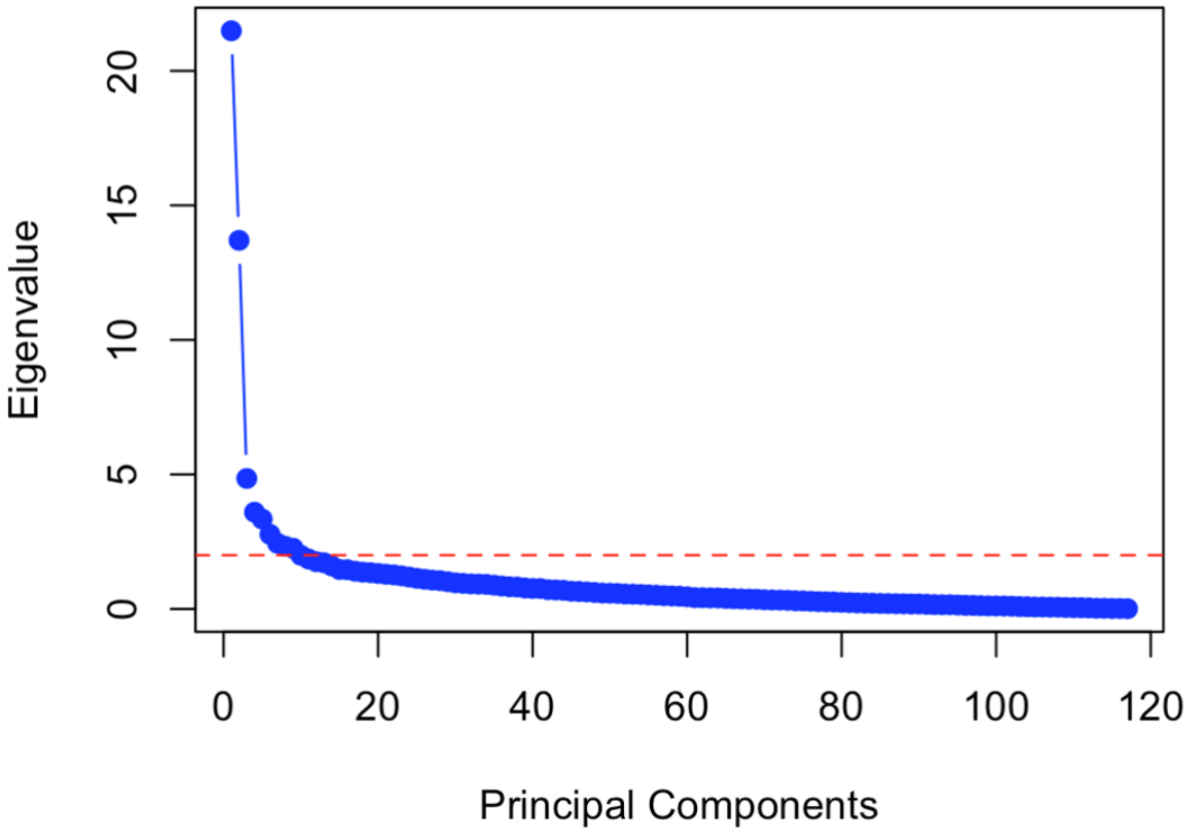


**Figure 5S | Scree plot of biomarkers' PC**
